# Estimating Preventable COVID-19 Infections Related to Elective Outpatient Surgery in Washington State: A Quantitative Model

**DOI:** 10.1101/2020.03.18.20037952

**Authors:** Yuemei Zhang, Sheng-Ru Cheng

**Author notes:** Corresponding author: Sheng-Ru Cheng, Corresponding author’s.

## Abstract

**Background:** As the number of suspected and confirmed COVID-19 cases in the US continues to rise, the US surgeon general, Centers for Disease Control and Prevention, and several specialty societies have issued recommendations to consider canceling elective surgeries. However, these recommendations have also faced controversy and opposition.

**Methods:** Using previously published information and publicly available data on COVID-19 infections, we calculated a transmission rate and generated a mathematical model to predict a lower bound for the number of healthcare-acquired COVID-19 infections that could be prevented by canceling or postponing elective outpatient surgeries in Washington state.

**Results:** Our model predicts that over the course of 30 days, at least 75.9 preventable patient infections and at least 69.3 preventable healthcare worker (HCW) infections would occur in WA state alone if elective outpatient procedures were to continue as usual.

**Conclusion:** Canceling elective outpatient surgeries during the COVID-19 pandemic could prevent a large number of patient and healthcare worker infections.

## Introduction

Despite its humble origins as a cluster of cases restricted to Wuhan, China in Nov. and Dec. of 2019, COVID-19 spread explosively across the globe and was officially declared a pandemic by the WHO on March 11, 2020.^1^ In the United States, the number of confirmed cases has spiked from just 1 case between Jan. 20, 2020 to 4661 confirmed positives and 85 deaths as of March 16, 2020.^2^ Washington state, the original epicenter of the US outbreak and the location of the first American case, has had 904 COVID19+ patients as of March 16, 2020.^3^ Given its rapid spread and 3.4% mortality rate,^4^ countries like Italy and China have been forced to ration limited healthcare resources, and there are concerns that the US may need to do so as well.^5^ Person-to-person transmission by asymptomatic individuals and pre-symptomatic individuals during the up-to-14 day incubation period^6^ may play a significant role in this pandemic.^7-10^ Infection transmission between COVID-19 patients and healthcare workers has also been documented.^11^

Given the current status of the COVID-19 outbreak, the US Surgeon General,^12^ Centers for Disease Control and Prevention (CDC),^13^ American College of Surgeons (ACS),^14^ American Society of Anesthesiologists (ASA), and Anesthesia Patient Safety Foundation (APSF)^15^ have recommended considering rescheduling or postponing some elective surgeries with the goal of conserving limited resources, such as ventilators and ICU beds, and mitigating the risk of “exposing other inpatients, outpatients, and health care providers to the risk of contracting COVID-19” from asymptomatic but infectious patients.^14^ However, the American Hospital Association, the Federation of American Hospitals, the Association of American Medical Colleges, and the Children’s Hospital Association have written a joint letter opposing the surgeon general’s advice.^12^ Multiple hospitals, including several major hospital systems in WA, are canceling or postponing elective surgery procedures,^16,17^ but there are other hospitals that have declared they will proceed with elective cases.^18^

The goal of this study is to provide a quantitative analysis and model for preventable COVID-19 infections from elective outpatient or ambulatory surgery cases. Our model can also be adapted to analyze COVID-19 transmission in other healthcare settings. Furthermore, given the controversy over the appropriate handling of elective surgical cases during this pandemic, we hope that our results may have a positive impact on health policy and public health. Given much of the uncertainty regarding the pathophysiology and epidemiology of COVID-19, and the potential policy implications of our results, we chose to focus on lower bounds for preventable infections instead of upper bounds.

We are excluding symptomatic COVID19+ patients from our model because their elective surgeries would likely be postponed or canceled due to the significantly increased risk of postoperative pulmonary complications if a surgical patient had a recent acute respiratory infection.^19^ Thus, our elective surgery patient population only includes uninfected individuals and asymptomatic or pre-symptomatic individuals (whose COVID-19 status would not be discovered given current testing limitations). Since COVID-19 would not be suspected in these patients, healthcare workers interacting with them typically would not use the level of personal protective equipment (PPE) or precautions necessary to prevent COVID-19 transmission, especially if there were also restrictions due to PPE shortages within the clinical institution.

## Methods

### Data sourcing

Since state-specific data on the daily volume of elective outpatient surgeries performed was not publicly available, we had to derive this number from available national data. The elective surgery population is estimated using data from the National Health Statistics Reports on Ambulatory Surgery Data in 2010. According to the report, an estimated 48 million elective ambulatory surgeries occur annually in the US.^20^ This number excludes outpatient elective surgeries performed in hospitals, so the actual daily volume of all outpatient elective surgeries in WA is likely higher. Since elective cases are not performed daily but every center or healthcare institution has different holiday schedules and policies, we divided this number by 365 days/year for a lower bound of 131,506.85 cases per day nationally. To simplify the calculation for the estimated number of elective outpatient cases in WA, we assumed that the case number was directly proportional to population. We divided 131,506.85 daily cases by the US population estimate of 328 million,^21^ then multiplied the quotient by WA’s population of 7.6 million,^22^ to arrive at approximately 3047 elective outpatient surgeries per day in Washington state.

In order to predict the lower bounds for the number of preventable patient and healthcare worker infections, we decided to minimize the number of unique healthcare workers (HCW) that patients would interact with in an elective outpatient setting. We derived this number for two reasons: 1. The number of clinical staff involved specifically in perioperative care for outpatient elective surgeries is unavailable and 2. Given the current state of the pandemic, institutions may attempt to minimize the numbers of non-essential staff involved in the care of each patient in order to reduce the risks of COVID-19 transmission. At minimum, each patient must interact with 4.5 HCW: one anesthesiologist, one surgeon or proceduralist, one circulator, one scrub technician, and 0.5 pre-operative / Post-Anesthesia Care Unit (PACU) nurses for both pre-op and post-op care, since the PACU nursing ratio is usually 1 nurse to 2 patients and the same nurse can care for a patient during pre-op and post-op. Note that the actual number of HCW that patients will interact with can often be higher. The number of patients that each set of perioperative staff works with varies depending on the length of surgery and scheduling preferences. Based on the clinical experiences of one of our authors, we will use the assumption that each HCW is responsible for an average of 5 cases or 5 unique patients. Thus, we came up with the ratio of 4.5 HCW / 5 patients, or 0.9 HCW/patient. Given our estimate of 3047 ambulatory surgery patients per day, this approximates 2742 HCWs involved in ambulatory surgery care per day.

### Initial conditions

**Fig 1.**
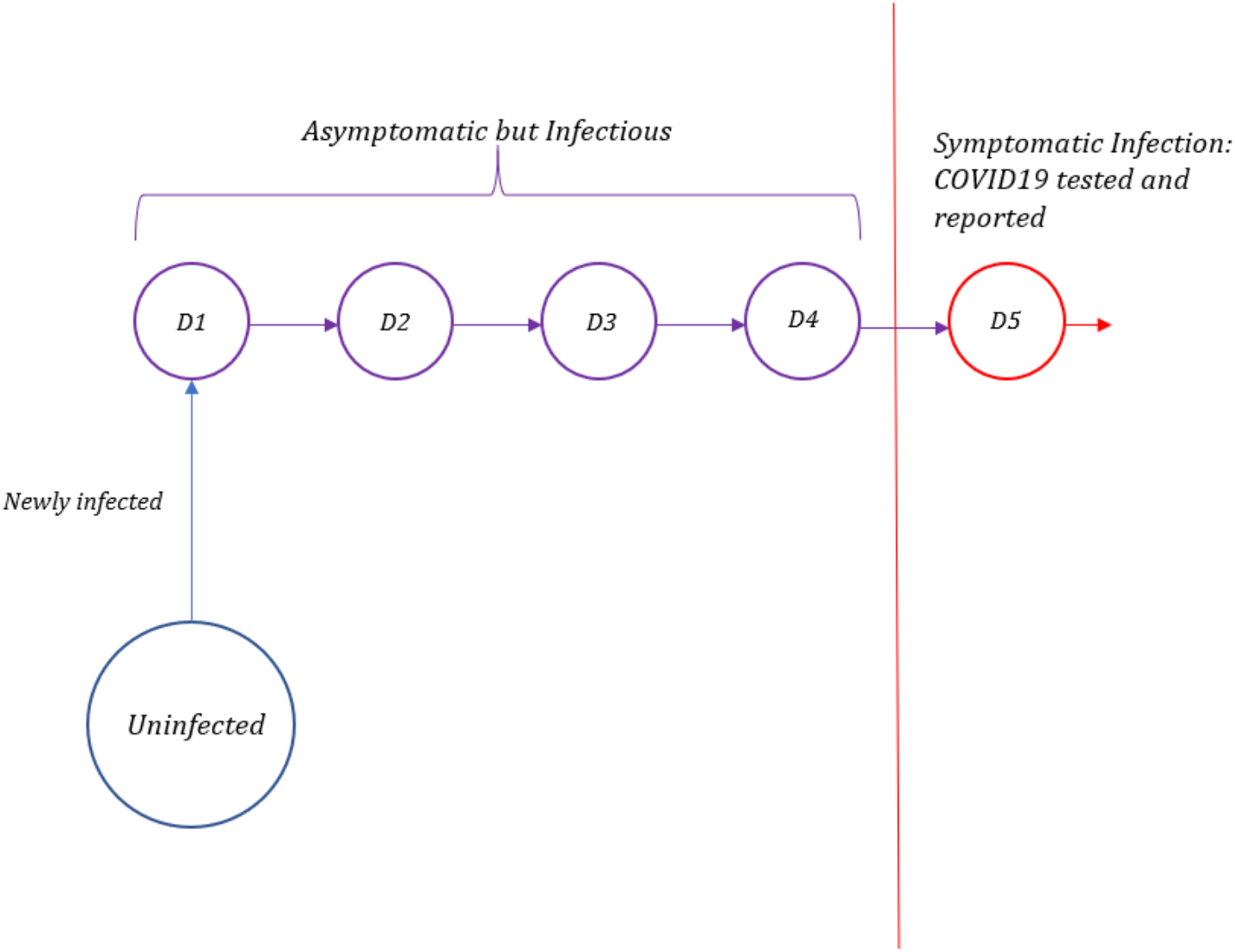
Timeline of Infection for Confirmed COVID-19 Cases. After infection, the individual can transmit the infection to others but does not become symptomatic until day 5, at which point they become eligible for COVID-19 testing and their infection is discovered and reported.

Due to the incubation period of the virus,^6^ coupled with the current resource limitations in the US, COVID-19 infections will not be detected until symptoms become evident. To estimate the asymptomatic infected population, we looked at confirmed COVID-19 cases and back-calculated the population count that would have likely been in the pre-symptomatic incubation phase on previous dates. Based on recently published studies, the average incubation period of COVID-19 is around 5 to 6 days.^23-25^ For this model, we used the shorter incubation period of 5 days, meaning that symptoms begin on day 5. This means that, for any time t, the number of asymptomatic but infected individuals can be estimated using the sum of new infections that were confirmed on t + 1 to t + 4 as follows:

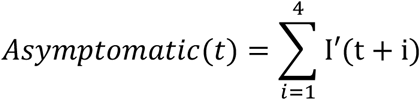

In other words, if someone is symptomatic and confirmed to be COVID19+ on any of the days between t+1 to t+4, then s/he was infected but asymptomatic on day t. Using data for Washington through March 17, 2020, we were able to calculate that there were at least 447 asymptomatic cases on March 13, 2020. However, this likely underestimates the actual prevalence of asymptomatic cases because this only includes the infected individuals that eventually tested positive, excluding those who ultimately never developed symptoms or only developed mild symptoms, those who developed severe symptoms but were never tested, and those who were tested but had a false negative result. Furthermore, the number of COVID-19 infections in the US and in WA have been steadily increasing over time.^2, 3^ News reports that the virus is thought to have been circulating within communities for weeks prior to the outbreak also support the idea that this number underestimates the actual prevalence of asymptomatic cases.^26^

Next, we needed to determine the ratio of asymptomatic uninfected people to uninfected people in the general population. We subtracted the confirmed infections on March 13, 2020 and the asymptomatic infected population on that day from the total population of WA in order to determine the uninfected population.

We assume that since the majority of patients and HCWs reside in WA, their infection statuses would initially also be representative of that of the general WA population. Thus we multiplied our ratio with 3047 total patients per day and 2742 total HCW to arrive at the initial values of 0.18 asymptomatic infected patients per day, 3046.82 uninfected patients per day, 0.16 asymptomatic infected HCW, and 2741.84 uninfected HCW. Asymptomatic infected HCW were further subdivided into groups based on how long they had been infected. Because asymptomatic COVID19+ individuals would remain in the workforce, we included infected HCW in the perioperative workforce for days 1-4 of their infections (during which time they could also infect other HCW and patients), and then removed them from the workforce once they reached day 5 and displayed symptoms. For our initial conditions, we divided asymptomatic infected HCW evenly into 4 groups for HCW on day 1 of infection (D1), day 2 of infection (D2), day 3 of infection (D3), and day 4 of infection (D4).

### Transmission rate

To investigate the number of preventable infections of healthcare workers from asymptomatic infected patients, we used a simple logistic model of transmission:

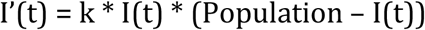

In this equation, k is the transmission constant, I’(t) is the rate of change of infected population, and I(t) represents total infected population, including the asymptomatic infected population. Since I’(t) is the rate of change of the infected population, it can be observed that the number of total infected population at a discrete time t + 1 is calculated as

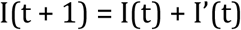

To calculate the transmission constant, we rearrange the previous equations to the following

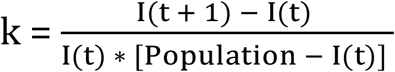

Since we are interested in the total infection spread, data for some known infected population, both symptomatic and asymptomatic, is required. For this, we used data extracted from the Diamond Princess cruise ship,^27^ since the close quarters approximate the perioperative setting. Due to the isolated nature of the ship, health officials were able to test everyone onboard the cruise ship, even if there are no symptoms evident. Using the data at hand and the equation above, we can readily determine the transmission constant by dividing the number of new cases at time t + 1 (with time measured in days) by the product of infected population at time t and the uninfected population at time t, which we calculated to be an average of k = 1.219e-4.

To calculate the number of preventable patient-to-HCW infections, HCW-to-patient infections, and HCW-to-HCW infections, we adapted the logistic model to

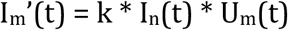

where I_m_’(t) refers to the new infections of a population *m*, k is the transmission constant, I_n_(t) refers to asymptomatically infected individuals of the group *n* transmitting the virus, and U_m_(t) refers to uninfected individuals of the group *m* that is being newly infected. For instance, if I_m_’(t) represents new HCW-to-patient infections, then I_n_(t) would represent asymptomatically infected HCW, and U_m_(t) would represent uninfected patients showing up for surgery. These calculations would be repeated for every day in our model. Since HCW and their patients interact much more closely with one another than they would with members of the general population outside this relationship, and we assume patients and HCW are following infection prevention guidelines such as social distancing appropriately,^28^ we will assume the likelihood of either a patient or a HCW becoming infected with COVID-19 from outside the clinical setting is negligible in comparison. Because patients do not freely interact with one another in pre-op, the OR, or the PACU, we assume patient-to-patient transmission is negligible in comparison to the other types of healthcare-associated transmission.

By definition, outpatient surgery means that patients leave the institution each day and a new batch of patients with characteristics representative of the general population would arrive each day. Although in real life, complications can occur that necessitate inpatient stays following outpatient surgery, for simplicity, we did not include that possibility in our model. Therefore, the starting numbers of uninfected patients and asymptomatic infected patients that we used for our calculations stayed constant.

On the other hand, since HCW were unlikely to have significant changes in their employment in the time period we were modeling, we designed a Markov chain to track their infection timelines. New HCW infections comprised the D1 group for the following day, and HCW in D1 would get changed to D2 the following day, HCW in D2 would get changed to D3 the following day, so on and so forth.

**Figure 2.**
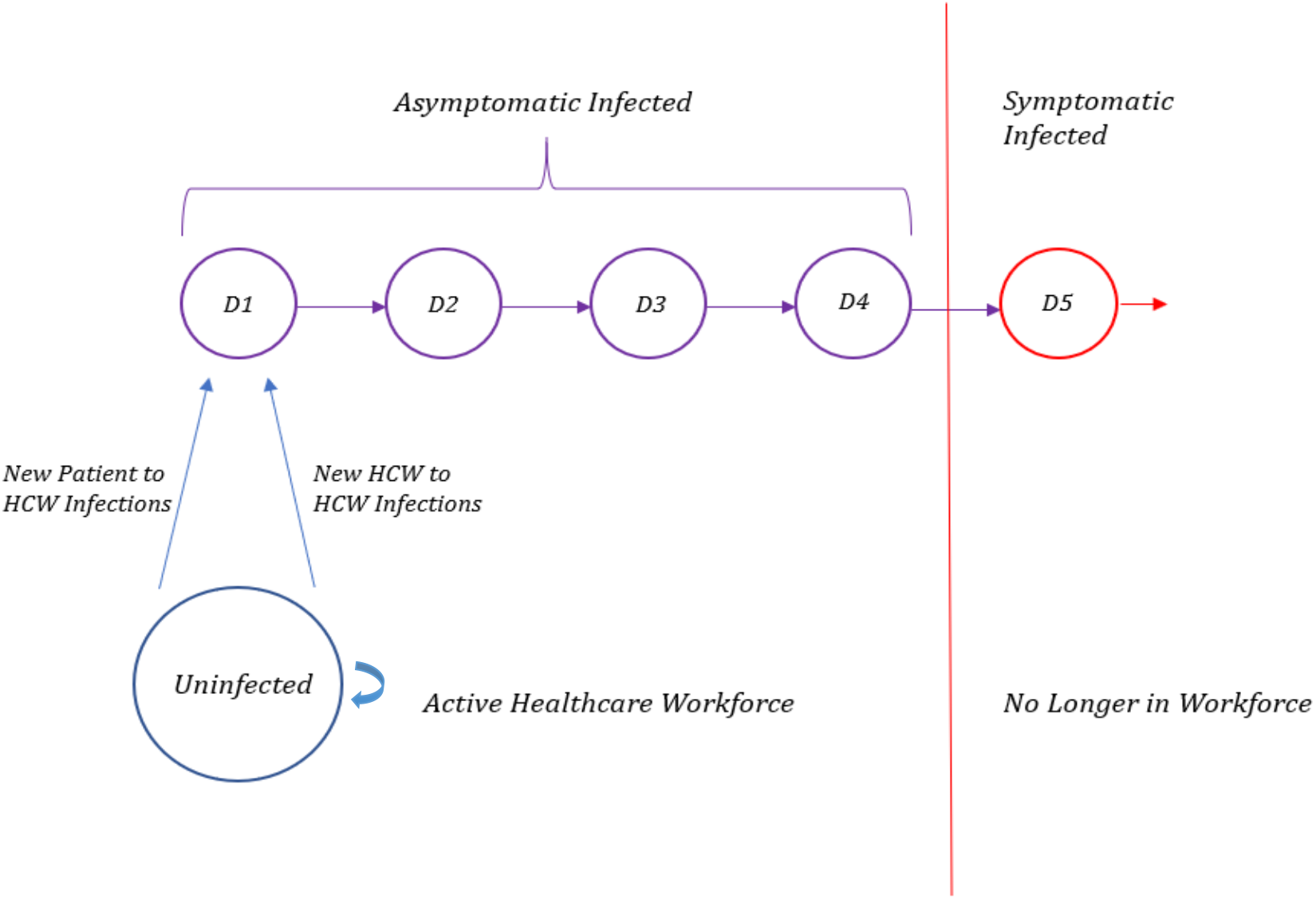
Markov Chain for Healthcare Workers. Healthcare workers (HCW) who are uninfected on any given day can either stay uninfected or become newly infected, at which point they would proceed to day 1 of infection the next day. Individuals who are infected will proceed to the next day of infection with each passing day. On days 1-4 of infection, infected HCW are asymptomatic and therefore continue to fully participate in the workforce, exposing other individuals to the risk of COVID-19 infection. On day 5 of infection, infected individuals begin showing symptoms, at which point they may no longer participate in the perioperative workforce.

## Results

### Preventable patient infections

Our model predicts that over the course of 30 days, 91,410 outpatients had elective surgery, and at least 75.9 of them developed preventable infections in WA state alone if elective outpatient procedures were to continue as usual.

### Preventable healthcare worker infections

Based on our model, over the course of 30 days, at least 69.3 preventable HCW infections would occur in WA state alone if elective outpatient procedures were to continue as usual. This represents approximately 2.5% of the initial perioperative workforce in our model. Of those infections, 1.8 can be attributed to patient-to-HCW transmission and 67.5 can be attributed to HCW-to-HCW transmission.

**Figure 3.**
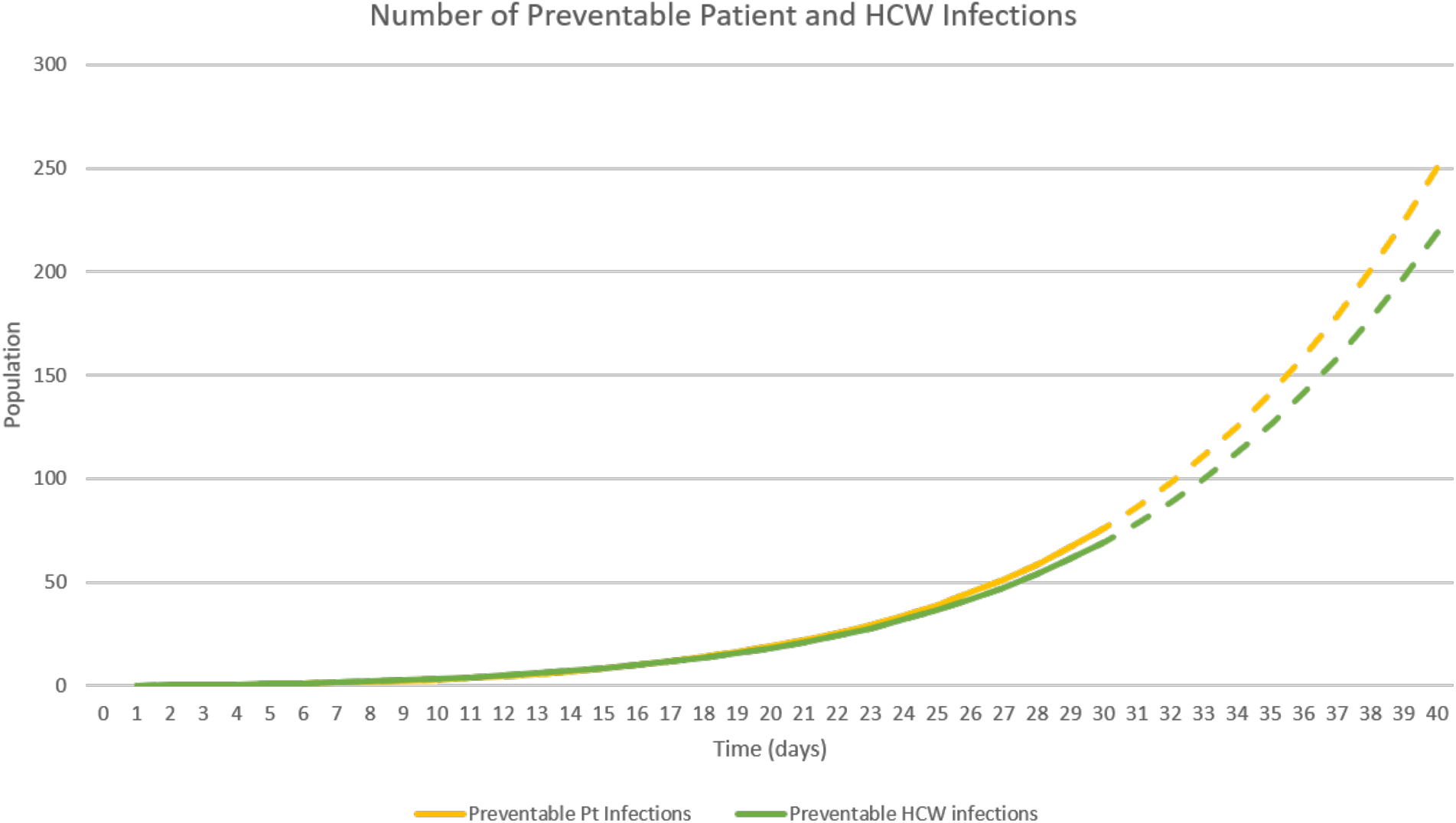
Number of Preventable Patient and HCW Infections. In the early phase of the pandemic, preventable patient infections (yellow line) and preventable HCW infections (green line) exhibit exponential growth, reaching a cumulative number of 75.9 preventable patient infections and 69.3 preventable HCW infections by day 30 attributable to outpatient elective surgery. The dashed lines represents projections if the same surgical volume were to continue, but in reality that is unlikely to happen given that HCW staffing would become an issue and start to limit case volume.

## Discussion

This model demonstrates that a substantial number of potential COVID-19 infections in both patients and HCW can be prevented by cancelling elective outpatient surgeries during this pandemic. In combination with the concern that we may not have enough healthcare resources for patients who are being admitted for COVID-19 symptoms, it appears that postponing elective surgeries may be an appropriate consideration.

Compared to the confirmed number of 904 positive COVID-19 cases in WA^3^ and 4661 confirmed positive cases in the entire US, a total of 145.2 preventable infections (69.3 HCW and 75.9 patient infections) over the course of a month is quite significant. While an argument can be made that a portion of these infections would have occurred regardless in non-healthcare settings, initiatives to limit public transmission would likely reduce that number. However, looking at the rapid growth of COVID19+ cases in the US, from 1 case on Feb. 29, 2020 to 4661 confirmed cases just 17 days later,^2^ it nevertheless should be noted that any mitigation should be implemented given the infectious nature of the virus.

Due to the current state of COVID-19 testing, US statistics on confirmed COVID-19 cases may not be the most reliable, either. Per CDC guidelines that were last updated March 9, 2020, laboratory testing for COVID-19 is only indicated for individuals who both develop respiratory symptoms consistent with COVID-19 and meet additional criteria, such as being hospitalized, having certain comorbidities, and/or having contact with suspected COVID19+ individuals.^29^ However, many COVID19+ individuals may be asymptomatic or only have mild symptoms.^30^ In addition, COVID-19 testing shortages may make the US statistics on COVID-19 cases less reliable.^31^ It is possible that many of our predicted new infections would not qualify for lab testing per the CDC’s guidelines or would not be able to access it, and therefore would not be included in COVID-19 case counts.

Of note, the majority of new infections are transmitted by asymptomatic infected HCW, not by patients. By exclusively examining outpatient surgeries, we have a constant flow of patients in and out of the system, whereas the HCW stay at the surgical center or hospital for much longer periods of time, and the proportion of asymptomatic infected HCW that patients interact with accumulates. While we did not look at other healthcare settings, this seems to suggest that minimizing the risk of COVID-19 infection to HCW in general may be important to preventing hospital-acquired COVID-19 infections in patients as well.

Given the uncertainty and unavailable data regarding COVID-19, some of the numbers and factual assumptions in this model may be incorrect, which could affect the model’s predictions. To simplify calculations, this model assumes that COVID-19 infections are spread homogenously throughout the state, that healthcare workers freely interact with patients and all other healthcare workers, and does not take into account individual variation in incubation times. Ultimately, this model is intended to be a tool and an approximation, and it can be adapted to different healthcare settings or regions by changing the starting conditions.

## Data Availability

The data that support the findings of this study are openly available reference number [3,23] and in the links provided.

https://github.com/CSSEGISandData/COVID-19

